# Protocol for the development of an artificial intelligence extension to the Consolidated Health Economic Evaluation Reporting Standards (CHEERS) 2022

**DOI:** 10.1101/2023.05.31.23290788

**Authors:** Claire Hawksworth, Jamie Elvidge, Saskia Knies, Antal Zemplenyi, Zsuzsanna Petykó, Pekka Siirtola, Gunjan Chandra, Divya Srivastava, Alastair Denniston, Anastasia Chalkidou, Julien Delaye, Petros Nousios, Manuel Gomes, Tuba Saygin Avsar, Junfeng Wang, Stavros Petrou, Dalia Dawoud

## Abstract

1.2.

**Introduction:** AI interventions for health care are on the rise. Decisions about coverage and reimbursement are often informed by Health Technology Assessment (HTA) bodies, who rely on Health Economic Evaluations (HEEs) to estimate the value for money (cost effectiveness) of interventions. Transparent reporting of HEEs ensures they can be used for decision making. Reporting guidance exists to support this, such as the Consolidated Health Economic Reporting Standards (CHEERS) checklist. We aim to identify consensus about specific items should be reported by HEEs that evaluate AI interventions and, if such items are identified, to develop them into an extension to CHEERS: “CHEERS-AI”.

**Methods and analysis:** The project will have 4 phases:

- Phase 1 is a literature review to help identify potential AI-related reporting items.
- Phase 2 commences a Delphi process, with a series of surveys to elicit the importance of the potential AI-related reporting items.
- Phase 3 is a consensus-generation meeting to agree on the final extension items.
- Phase 4 is dissemination of the project’s outputs.

**Ethics and dissemination:** This study has received ethical approval from Newcastle University Ethics Committee (reference: 28568/2022). The findings will be available in as an open access article and disseminated through blogs, newsletters, and presentations.

**Funding statement:** This study is supported by the Next Generation Health Technology Assessment (HTx) project. The HTx project has received funding from the European Union’s Horizon 2020 research and innovation programme under grant agreement Nº 825162. This dissemination reflects only the views of the authors and the Commission is not responsible for any use that may be made of the information it contains.

## 2. Introduction

In recent times there has been a rapid increase in the development of technologies with an artificial intelligence (AI) component for health care interventions. This is evidenced in the number of approvals given by regulatory bodies. Between 1997 and 2021, the Food and Drug Administration in the United States approved 350 AI technologies with 91% of them approved since 2015 (1). In 2021, the European Medicines Agency (EMA) led a report on behalf of the International Coalition of Medicines Regulatory Authorities documenting a horizon scanning exercise in AI and highlighting regulatory challenges (2). This was in response to these new technologies increasingly challenging regulatory frameworks and a need for recommendations on how to adapt them.

AI is a broad term to encompass iterative, ‘learning’ algorithms that use data and high computing power to make interpretations, predictions or decisions (2). Some AI technologies are fixed, and others are adaptive. Various subsets of AI, such as machine learning (ML), are being used throughout the drug discovery process for target validation, identification of biomarkers, and analysis of clinical trial data (3). As well as assisting with the drug development process, AI is also featuring in the end product, and it is these health technologies that are the focus of this paper. Examples of AI health interventions include systems for screening and triage, diagnosis, prognosis, decision support, and treatment recommendation (4,5).

To ensure their appropriate use in healthcare pathways, we need to understand what benefits new AI technologies bring, and at what cost. There are established methods to do this for pharmacological and diagnostic interventions, but AI algorithms may be distinct from more traditional interventions in numerous challenging ways. Firstly, they have the potential to learn over time, meaning the relationship between intervention and outcome may not be fixed. This has implications when considering future benefits, such as choosing a suitable method or assumption for long-term treatment outcomes. We often see an assumption that the treatment effect of a medicine wanes over time, but how might healthcare decision makers appropriately value on an AI intervention that might get *more* effective over time? Secondly, the user is most often a health care professional rather than a patient, and the degree to which the clinician employs the results of the AI intervention may vary, particularly when its purpose is a decision-support tool. Thirdly, trial data normally underpin a health technology assessment (HTA) and reimbursement decisions. However, to date, AI technologies have not typically been subjected to interventional trials, meaning various data sources or assumptions will be required to inform a value assessment. Although randomised controlled trials (RCTs) are increasingly being conducted to evaluate the clinical efficacy of interventions with an AI component, there are concerns relating to their design and reporting. To try to address these concerns, AI extensions to reporting checklists have been developed; for example, for protocols (SPIRIT-AI) (5) and trials (CONSORT-AI) (4).

In addition, the ways in which AI-based intervention are developed arguably create an extra, inherent layer of uncertainty. Their function and attainment depend on the data sets used to train and validate their underlying algorithms. This development, or learning, step precedes any study of efficacy relative to the standard of care, which tends to be the primary source of potential uncertainty for more traditional interventions.

Health economic evaluations (HEEs) assessing the cost effectiveness of health interventions are often used by HTA bodies to make their reimbursement recommendations. HTA bodies will increasingly be expected to assess the value of health technologies that use AI. For example, the National Institute of Health and Care Excellence (NICE) in the UK recently updated their Evidence Standards Framework to reflect and include adaptive AI and data-driven technologies (6,7). The usefulness of a published HEE to decision makers depends on how well it is conducted and reported. Reporting guidelines can improve their transparency and completeness. A prominent HEE reporting checklist is the Consolidated Health Economic Evaluation Reporting Standards (CHEERS) statement. It was originally published in 2013 to help authors accurately report details of the HEE, including the health intervention, what was being compared and in what context, how the evaluation was undertaken, and what the findings were (8). This checklist outlined minimum reporting standards and the increased transparency allows decision-makers such as HTA bodies and payers to judge the quality and appropriateness of the HEE for their decision problem, facilitating trust in the results. The CHEERS statement was updated in 2022 (9) and now comprises a 28-item checklist including methodological approach, data identification, model inputs, assumptions, uncertainty analysis, and conflicts of interest.

CHEERS 2022 does not include any reporting items that are specific to potential AI components of an intervention, but the authors of CHEERS 2022 explicitly “encourage those who see opportunities to expand CHEERS 2022 items or create additional reporting guidance that provides clarification in specific areas to work with members of the CHEERS Task Force to develop CHEERS extensions in these areas”. As noted above, extensions for AI health interventions have already been developed for other checklists, demonstrating a system wide need and motivation for improving best practice around data collection and transparency. Including AI-specific items in the reporting of HEEs is a logical step to contribute to this standard setting for AI interventions. It will help to ensures that all relevant information required for decision-making is available to decision-makers.

## 3. Methods

Our research approach was guided by the EQUATOR (Enhancing the QUAlity and Transparency Of health Research) Network’s recommended steps for developing a health research reporting guideline (10) and methods used to develop other related extensions (CHEERS 2022, CONSORT-AI and SPIRIT-AI). The guideline extension is registered on the EQUATOR Network website (11). The structure and writing of this protocol were guided by the recently published protocol for the SPIRIT-SURROGATE and CONSORT-SURROGATE extensions (12).

A project management group led by NICE is organising and conducting the project with oversight from a Steering Group. The Steering Group is a multi-disciplinary and international group with representation from University of Oulu, Finland; Zorginstituut Nederland (National Health Care Institute) and Utrecht University, the Netherlands; Syreon Research Institute, Hungary; Tandvårds-och läkemedelsf□rmånsverket (The Dental and Pharmaceutical Benefits Agency), Sweden; and The London School of Economics and Political Science, the University of Birmingham, University College London and University of Oxford, UK. The Steering Group includes a representative from the CHEERS Task Force to provide expert input. The Steering Group was formed in December 2022.

This study has been supported by Next Generation Health Technology Assessment (<u>HTx</u>), which is a Horizon 2020 project supported by the European Union, lasting for 5 years from January 2019. Its main aim is to create a framework for the next generation of HTA to support patient-centred, societally oriented, real-time decision making on access to and reimbursement for health technologies throughout Europe.

### 5.1. Phase 1: Systematic literature review

We conducted a systematic literature review in the summer of 2022 to assess the methodological and reporting quality of HEEs of AI-based technologies. This updated a previously published review by Voets et al (13). Our search was performed on 17^th^ June 2022 and found 21 HEE studies published in the preceding 15 months. This review was used to identify potential AI-extension items. Members of the Steering Group were also able to contribute potential AI-extension items, based on their knowledge and experience of AI, HEE, and reporting guideline development. This led to a ‘long-list’ of potential items for an AI extension to CHEERS 2022. The review and Steering Group also helped to identify subject matter experts who could participate in the Delphi study. This group are referred to as the Expert Panel (EP).

### 5.2. Phase 2: Consensus-generation surveys (Delphi process)

This phase will involve participants rating long-list candidate items generated in phase 1 and suggesting additional items not included in the long-list. There will also be the opportunity to revise wording for the items. Proposed timelines are to open the first survey round in May 2023 to coincide with the ISPOR 2023 conference in Boston, US. The consensus process will dictate the number of necessary survey rounds, but it is anticipated that the whole project will complete in during 2023.

#### 5.2.1. Survey design and setting

This methodology follows that used for the development of CHEERS 2022 (9). CHEERS 2022 employed a modified Delphi process. Delphi is a widely recognised and used method for consensus-building and revolves around the following key steps: identification of factors, anonymous surveys among subject matter experts to elicit importance, integration and controlled feedback and presentation of aggregated data at consensus meetings (14). The survey will be developed in <u>Snap Surveys</u> software.

We will conduct a minimum of 2 survey rounds and will consider additional rounds if necessary. This approach has been taken for other guideline extensions (4,5,9,12).

#### 5.2.2. Sample size, recruitment, and inclusion criteria

We will recruit an EP representing the following key stakeholder groups: health economists, AI methodologists and academics, industry, policy makers, HTA experts, ethicists, patient representatives, journal editors, healthcare professionals, payers, and research funders. This is consistent with the EQUATOR Network’s guidance (10) and groups who participated in related extension. We anticipate inviting over 100 EP members.

Our approach to recruit EP members involves a multi-faceted approach. The Steering Group will identify participants. This purposive sampling will utilise a snowball sampling method where invited participants will be allowed to invite additional participants, meaning the total number of survey recipients should far exceed the those identified by the Steering Group. The survey will elicit the profession of the recipient to ensure that all respondents are part of one or more of the key stakeholder groups. We will also approach relevant professional groups such as the ISPOR Machine Learning Task Force. We will utilise authors identified in the phase 1 as another source of potential participants and will coordinate completions of the survey with the ISPOR conference, taking place in May 2023 in Boston, US. All EP members will be sent an introductory email and participant information sheet.

We will collect descriptive demographic data at the start of the survey, including stakeholder group, country of work and years of relevant experience to indicate understanding of AI in healthcare and HEE. Inclusion criteria are the key stakeholder groups previously specified. There are no exclusion criteria, but this targeted recruitment should result in identification of suitable EP members.

There is guidance on the minimum number of survey responses to allow statistical rigour, with 30 commonly cited (15). By identifying and inviting over 100 experts to participate, we will allow sufficient headroom for non-response and attrition between survey rounds.

#### 5.2.3. Data collection, analysis, and consensus definition

The survey will be developed in consultation with the Steering Group, including a pilot prior to the launch to ensure usability. All participants will be sent a link to the survey which will start with study information and a tick box for consent. The first survey will be open for a 3-week window, commencing May 2023. The second survey will be sent approximately 4 weeks after closure of the first. Response rates will be monitored during the survey window and email reminders sent to participants to increase response rates. Records will be kept of the number approached, and non-responses.

The EP will be asked to vote on the relevance of candidate items when reporting a HEE of an AI-based intervention. They will be asked to use a 9-point Likert rating scale, consistent with CHEERS 2022 and other reporting extensions. A ‘don’t know’ option will also be available for each item, in case a participant feels unable to provide a rating. Potential items will be grouped according to standard sections of HEEs (e.g., title, abstract, methods, discussion), and each item will have an accompanying definition and rationale for inclusion. We will employ the consensus definition used for CHEERS 2022 (see Figure 1). After survey round 1, any items that were scored lower than 7 by at least 70% of respondents will be excluded; that is, we will conclude that consensus has been reached that those items are not relevant reporting standards for HEEs of AI interventions. Those items will be ‘rejected’ and will not proceed to survey round 2 in their original form. Participants will have the opportunity to comment on the wording and propose new items. If suitable revised wording of original items has been proposed, then revised items may be included in survey round 2.

**Figure 1.**
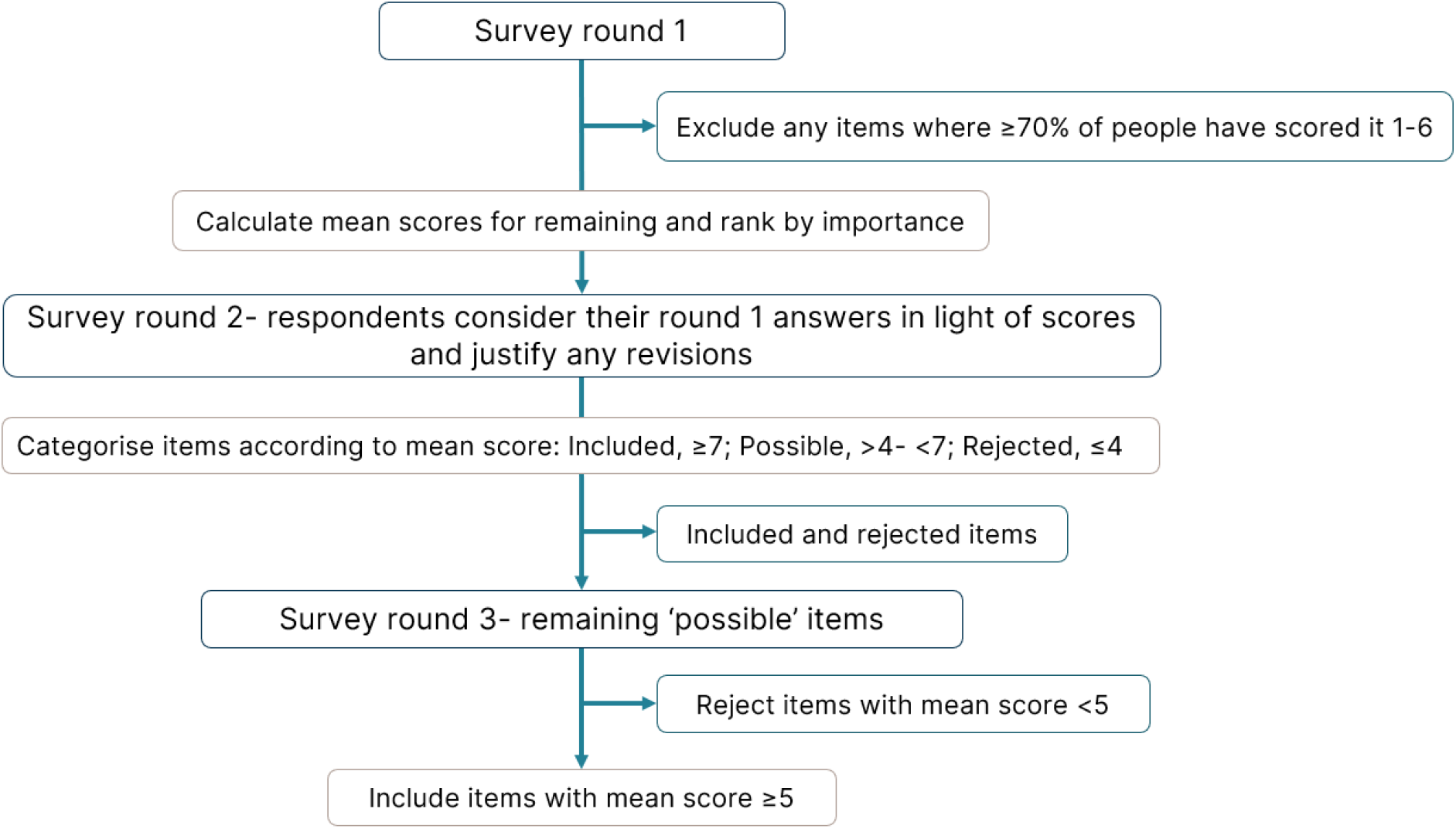
Flowchart showing the process for including and excluding items during the survey rounds.

Results from survey round 1 will be analysed in MS Excel to identify the mean scores and measure of agreement (proportion of scores 7 or higher). After excluding those meeting the exclusion threshold, the remaining items will be included in survey round 2. They will be presented in order of mean score and the measure of agreement will also be shown. Respondents will have the opportunity to consider their round 1 answers in light of the aggregate results and justify any revisions. Any new items that were suggested by respondents in survey one will also be voted on in the second survey round, along with proposed changes such a new wording or merging of items from free-text responses.

After survey round 2, results will be analysed in MS Excel, and items with a mean score of 4 or less will be categorised as ‘rejected’ (consensus reached). Items with mean scores of 7 or higher will be grouped as ‘included’ (consensus reached). Items with mean scores above 4 but less than 7 will be grouped as ‘possible’ (consensus not reached). Any such items will proceed to survey round 3, presented in order of importance (mean score), alongside a measure of agreement (proportion of scores 7 or higher). After this final survey round, items with a mean score of 5 or less will be ‘rejected’ (consensus reached). Items with a mean score above 5 will be ‘included’ (consensus reached). Therefore, consensus will be achieved for all items after survey round 3, unless participants provide substantial and conflicting free-text responses about the wording, or additional items. Any such items will proceed to a consensus meeting for resolution.

### 5.3. Phase 3: Consensus-generation meeting (Delphi process)

A consensus meeting will be held virtually, with the aim of concluding the final extension list. We will invite a purposive sample representative of the EP who completed every survey round. The EQUATOR Network has guidance on conducting face to face consensus meetings (10).

#### 5.3.1. Structure and participants

The COVID-19 pandemic has normalised virtual working and we propose to hold the meeting virtually. This is also advantageous in terms of maximising engagement and attendance from a range of geographical locations. The meeting length will be agreed after survey round 3, at which point the number of items that still haven’t achieved consensus (and therefore require extensive discussion) will be known. However, the meeting time will be sufficient to allow discussion time for all items. Other extensions have used meetings over two days (4,5,12).

At the end of survey round 2, participants will be invited to register their interest in attending the consensus meeting. The Steering Group will purposively select members from this pool considering the need to have an international multidisciplinary group of participants.

#### 5.3.2. Consensus procedure

Attendees at the meeting will ratify all items about which consensus was reached during the first 2 survey rounds. Items that proceeded to survey round 3 will be discussed more comprehensively at the meeting. Minor modifications to wording can be included by a simple 50% majority vote during the meeting.

The richest discussion will be for any items where consensus has not yet been reached by the end of the survey round 3. These will be any new items that were proposed in free-text responses during survey round 3, and any items that received extensive requests for modification in free-text responses (e.g., merging items), such that a simple 50% majority vote at the meeting would not be appropriate to support inclusion. For these significant modifications, a 70% majority vote during the meeting will be required to include the modified or new items.

Items that do not reach consensus (e.g., due to a large proportion of ‘don’t know’ or abstained votes) will be discussed further and voted on again, if appropriate, until consensus is reached or time runs out. The Steering Group will make final decisions soon after the consensus meeting on any outstanding items without consensus.

The consensus meeting will be recorded to ensure accurate recall, and minutes will be taken.

### 5.4. Phase 4: Knowledge translation

This phase includes all activities aiming to publish and publicise the extension. This objective will be integrated and considered throughout all stages of the project.

#### 5.4.1. Pilot testing and revision of final checklist

After the meeting we will conduct a pilot of the finalised extension with invited researchers to check clarity of wording and identify any challenges. These will be people whom we invited but were unable to join our Steering Group or who expressed an interest to join after survey round 1 had started. This exercise will inform writing of the explanation and elaboration documents.

#### 5.4.2. Publications

We aim to publish the CHEERS-AI extension in a high impact open access journal to maximise dissemination. We will also utilise the ISPOR CHEERS Task Force to help publicise the extension. We will seek the endorsement of the extension from journals and editorial groups.

#### 5.4.3. Partner and stakeholder engagement

The project is registered on the EQUATOR website. We aim to have the final extension published on the CHEERS website. The CHEERS statement is endorsed by ISPOR.

#### 5.4.4. Patient and public involvement

We have patient advocacy on the Steering Group led by EURORDIS (Rare Diseases Europe). They will ensure views and perspectives of patients and the public are represented at all stages. Patients and the public are also one of our key stakeholder groups to be represented on the Expert Panel and therefore will be involved in consensus building for the final checklist. EURORDIS and the NICE lead for patient and public involvement will be requested to advise on dissemination activities.

#### 5.4.5. Ethics and dissemination

An ethics application was submitted via the NICE ethical approval process. Ethics approval was received from Newcastle University Ethics Committee who are the awarding body (reference: 28568/2022).

Expert Panel members will be provided with a participant information sheet and will be asked to provide consent before completing the first survey. We will also obtain electronic written consent before the consensus meeting for participation and recording of the meeting. Participants will have a right to withdraw at any stage of the project. All data will be securely stored although we anticipate that it will not be highly sensitive. We will ask participants if they prefer to opt out of acknowledgement in any publications.

## Data Availability

The findings will be available in as an open access article and disseminated through blogs, newsletters, and presentations.

## Notes

### Competing Interest Statement

The authors have declared no competing interest.

### Author Declarations

Newcastle External Assessment Centre (EAC) of Newcastle University gave ethical approval for this work (reference: 28568/2022).

